# Prevalence of Herbal Medicine use among Pregnant Women Attending Antenatal care at Jimma University Medical Center, Jimma, Southwest Ethiopia

**DOI:** 10.1101/2022.11.16.22282405

**Authors:** Firafan Shuma, Damisie Bahiru, Dugasa Erko, Tamirat Tekassa

## Abstract

**Background:** The use of herbal medicine during pregnancy for different indications now a day becomes common across globally. Its use is increasing, especially in developing countries. It is estimated that 65%-80% of the population use herbal products

**Objectives:** This study aimed to assess the prevalence of herbal medicine use among pregnant women attending Antenatal care at Jimma University medical center, Southwestern Ethiopia.

**Methods:** A cross-sectional study design was employed to conduct the study. Data was collected by data collectors using data abstraction formats, then entered and analyzed using SPSS version 26.0. Frequency and percentage were computed as summary statistics.

**Results:** A total of 341 pregnant women attending antenatal care were enrolled in the study, of which 331 participated in the study. Out of 331 pregnant women who participated in the study, 3.93% of them practiced self-medication with herbal medicine during their current pregnancy.

**Conclusions:** The most frequently practiced herbal medicine among pregnant women attending antenatal care at JUMC were Ginger and Damakese with the most common clinical indication of vomiting and headache respectively. The source of information for the participant’s herbal medicine use were mostly family and friends.

## Introduction

Self-medication is defined as the administration of medication by a patient without a physician’s prescription or in a way that is not directed by a doctor. It is considered a form of self-care in which a patient takes on the burden of treating or preventing illness without the assistance of a doctor by using over-the-counter or prescription-only medications (POMs) [1].

Self-medication practices among pregnant women have continued to be the most public health concern globally due to incidences related to increased abortion, fetal malformation, and resistance to antimicrobials that were found to have significant associations [2].

Self-medication has been increasing in many developing and developed countries and there is growing concerned about self-medication during pregnancy in many low-resource countries [3-4]. Many nations in Sub-Saharan Africa (SSA) have the potential to benefit from traditional medicine in terms of both health and economics, particularly if regulatory frameworks are put in place to ensure its protection and application. The impact of traditional medicines (TM) on pregnancy is not well understood. The safety of TMs used during pregnancy has to be better understood to decrease maternal fatalities and problems associated with pregnancy [5].

In Ethiopia, the use of herbal medicine is relatively common and affects roughly half of pregnant women attending antenatal care. Ginger, damakasse, garlic, tenaadam, and eucalyptus were the herbal remedies that were most frequently used during pregnancy. These most popular plant species’ potential negative effects on the fetus during pregnancy are unknown. The study showed that about 152 (38.0%) pregnant women used HM. The most frequent disease for HM use was the common cold [4]. It is not feasible to completely rule out teratogenic effects due to the paucity of research on many medicinal plant species. Collaboration between traditional healers and medical experts to spread knowledge about the use of medicinal plants would promote healthier pregnancies and better maternal and newborn health [6].

There are not much data in Ethiopia and no evidence is available specifically in the current study area, despite the possible risks associated with pregnant women using self-medication. Therefore, this study aimed to assess the prevalence of herbal medicine use among pregnant women attending Antenatal care at Jimma University medical center, Southwestern Ethiopia. The most likely research questions used were: What is the pooled prevalence of herbal medicine use among pregnant women attending ANC care in Jimma University Medical center?

## Methods

### Study setting

The study was carried out at the Jimma University Medical Center in Jimma Town from January 20, 2022 to April 30, 2022. Jimma Town is 352 kilometers southwest of Addis Ababa. According to data from the Jimma Town Health Office, the town has an elevation between 1,750 and 2,000 meters above sea level, a temperature range between 20 and 30 degrees, and an average annual rainfall of between 800 and 2,500 millimeters. The estimated population of Jimma Town as of the year 2019 G.C. is 205,384. There are 104,745 ladies and 100,639 males among them. In addition, there are 7127 pregnant women, 33,744 children under the age of 5, and 45,452 women who are of reproductive age.

Ten government health facilities, including two hospitals, four health centers, and four health posts, make up the town’s health service.

### Study design

A Hospital-based cross-sectional study design was employed.

### Study population

All selected pregnant women from those attending Jimma University medical center for Antenatal service during the study period were considered as the study population.

### Eligibility Criteria

#### Inclusion criteria

Pregnant women attending Antenatal care at Jimma University medical center and volunteering to respond were included.

#### Exclusion criteria

Pregnant women with severe mentally ill, and also who were unwilling to respond were excluded.

### Sample size determination and sampling procedure

The sample size was determined using the single population proportion formula by considering the p-value of 69.8% from a study done in Nekemte Hospital and a 95 % confidence level and 0.05 margin of error are taken [7]. Thus, considering the total source population of N= 7,127, P=0.698, a confidence interval of 95% (α=0.05), and d=0.05, after using the correction formula and including a 10% contingency, the sample size was n= 341. But, only 331 pregnant women attending antenatal care participated in the current study. During the data collection, eligible pregnant women attending antenatal care were included in their visit to the Antenatal care unit until the estimated sample size was reached. To perform the study, systematic random sampling was used.

### Study variables

Self-medication of herbal medicine use was considered the dependent variable and Socio-demographic characteristics of study participants, Gestational age, and history of herbal medicine use were considered independent variables.

### Data collection process

Data were collected using an interviewer-administered questionnaire that was created and translated into English after being modified by relevant studies. The questionnaires ask about the respondent’s obstetric history, use of herbal medicines, and sociodemographic variables. It was employed following preliminary testing on 5% of the same source population that wasn’t included in the sample.

### Data processing and analysis

The data was cleaned and validated before being coded, inputted, and used in SPSS version 26.0. Analyses were done on the cleansed data. Frequency and percentage were computed as summary statistics.

### Data quality assurance

The questionnaires were translated into the local language, Amharic, and Afan Oromo and retranslated back to English for a constituency. Data was collected by data collectors.

### Ethical Considerations

This study was approved by the Jimma University Institute of Health Institutional Review Board (IRB) (ref. no. JUIH 143422/22) in Ethiopia. The hospital’s clinical director granted authorization for the investigation to proceed after receiving ethical clearance. A fact sheet was created and distributed to all potential study participants. All participants were made aware of the study’s objectives, and their participation was entirely voluntary. All participants provided verbal, informed permission. To maintain confidentiality, the participant’s name was not included in the questionnaire; instead, a medical record number was utilized.

### Dissemination plan

Jimma University; Institute of Health; school of pharmacy received the study’s findings in a presentation. JUMC, the Jimma Zone Health Bureau, and other stakeholders, including governmental and non-governmental groups, were also informed of the findings. Finally, attempts were made to publish in recognized publications journals.

### Definition of terms

#### Herbal medicines

Herbal medicines are defined by the World Health Organization (WHO) as plant-derived materials or products with therapeutic benefits, which contain either raw or processed ingredients from one or more plants [9].

#### Pregnancy

Pregnancy is a situation with many physiologic changes that cause plenty of pregnancy-relevant complications, including vomiting, nausea, heartburn, and constipation [8].

#### Self-medication

Self-medication is defined as the act of using medications by patients or individuals to treat self-diagnosed disorders or symptoms on their initiative without getting advice from a healthcare provider [10].

## Results

### Socio-demographic characteristics

The study comprised 341 pregnant women in total, and of those, 14.37, 52.49, 19.65, and 13.49% were between the ages of 18 and 23, 24-29, 30-35, and over 35, respectively (Fig. 1)

**Figure 1:**
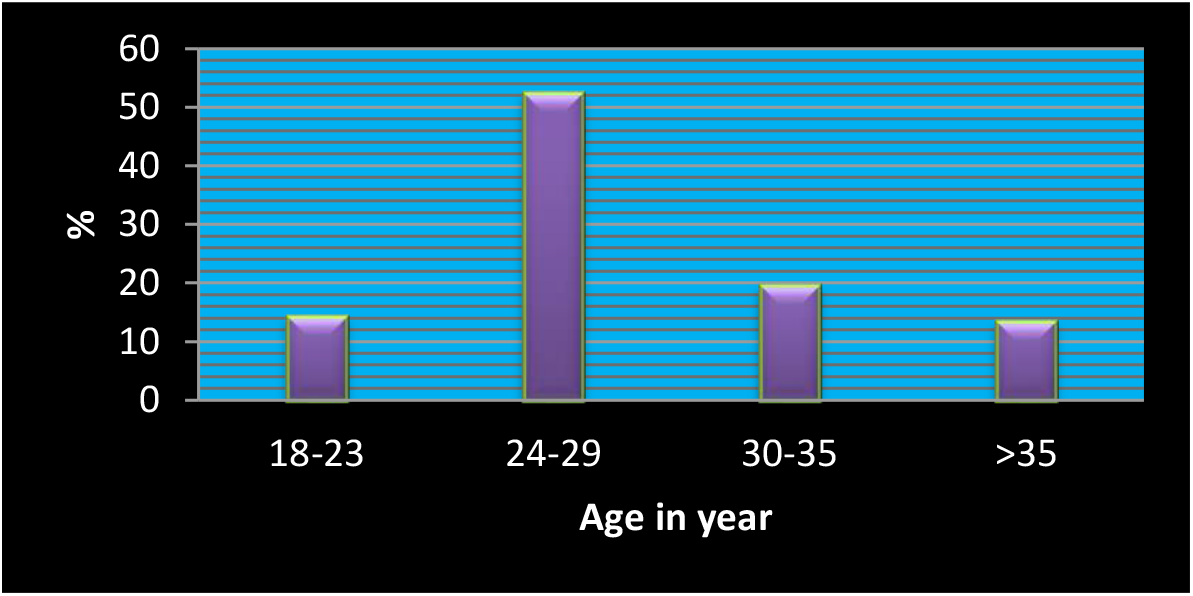
Percentage of respondents versus age

The majority of these pregnant women (93.55%) were married (Table 1). Regarding their occupation, housewives made up 67.45% of them, followed by farmers (24.93%), and 37.54% of them had low monthly incomes (less than 500 ETB). In addition, 271 (79.47%) of them were illiterate, 16.72% of them had attended elementary school (grades 1–8), and 265 (77.71%). The majority of the study participants were Oromo by their ethnicity and 265 (77.71%) of them were Muslims 82.99% (Table 1).

**Table 1.**
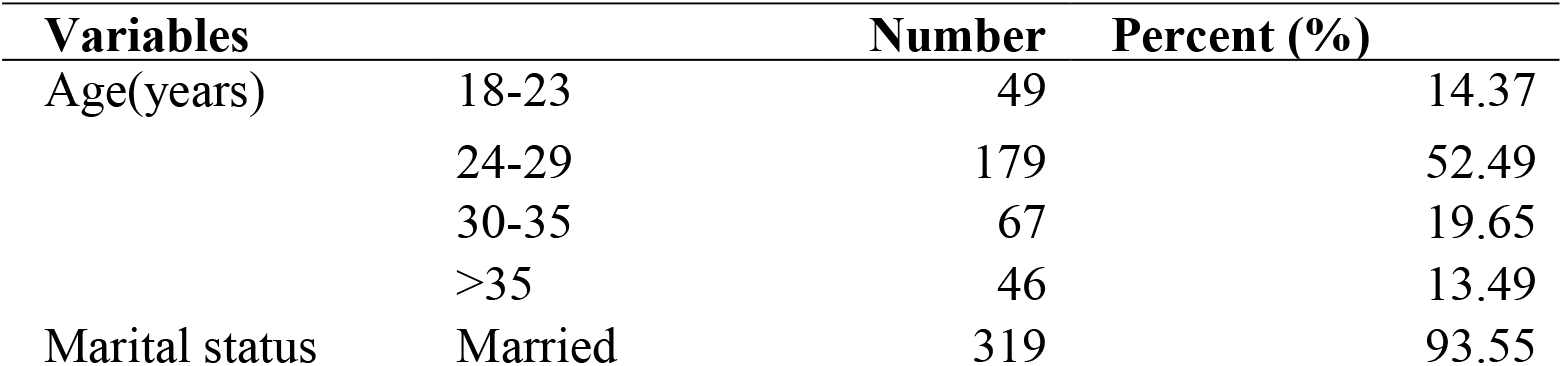
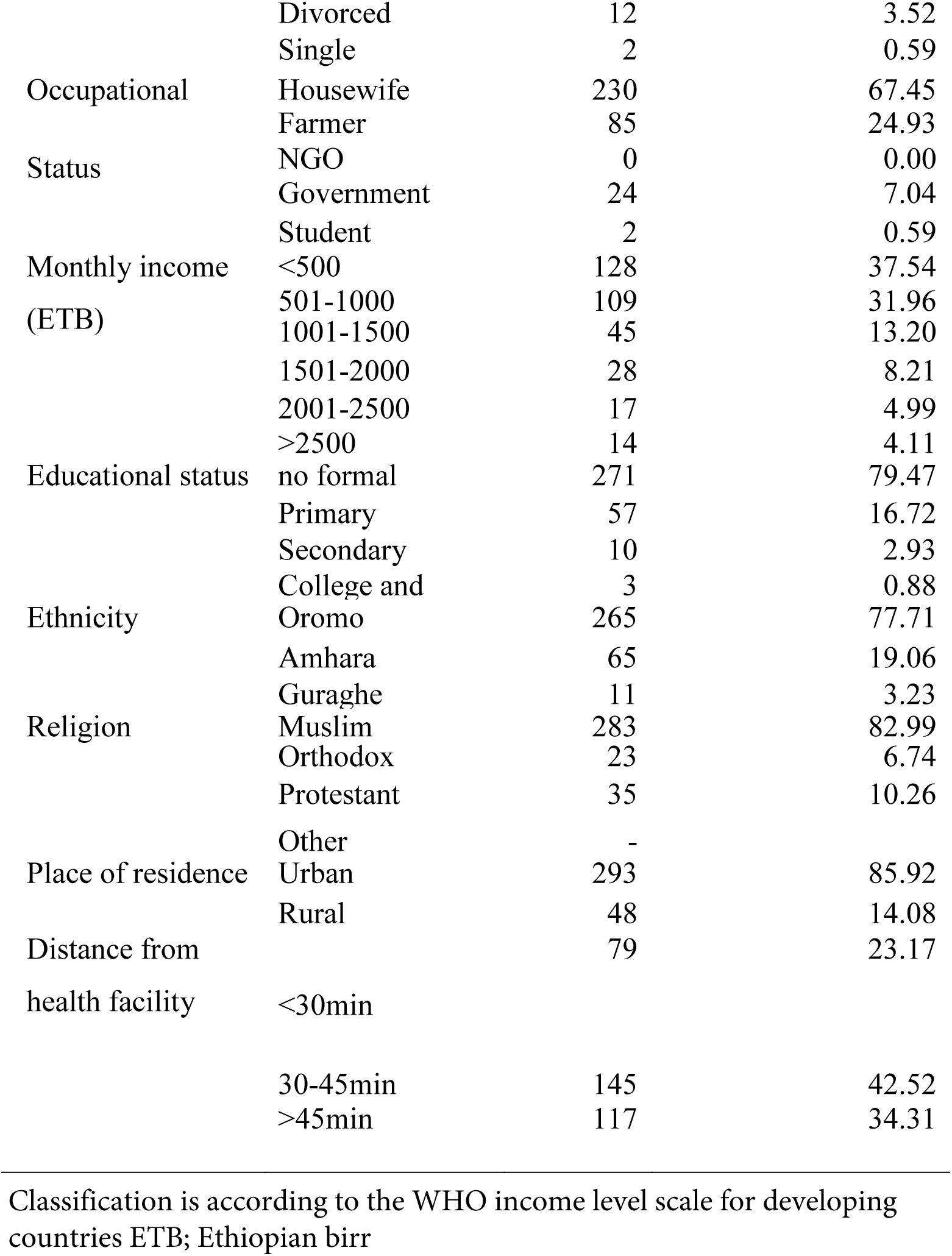
Socio-demographic characteristics of Pregnant women Attending ANC at JUMC

### Obstetric information of the respondents

Out of 331 pregnant women interviewed, 132 (39.88%) of them had two or fewer pregnancies, while 39 (16%) had four or more (Table 2). From a total of 331 pregnant women, 36 (10.88%) had more than three gravidities; 103 (33.30%) had two children; 18 (5.74%) had no children, and 4 (1.3%) had a history of past abortions (Table 2). Of the twenty-three expectant women who had previously had abortions, eleven (47.23%) had done so due to an undiagnosed sickness, and nine had done so as a result of using an unidentified medicine (Table 2).

**Table 2.**
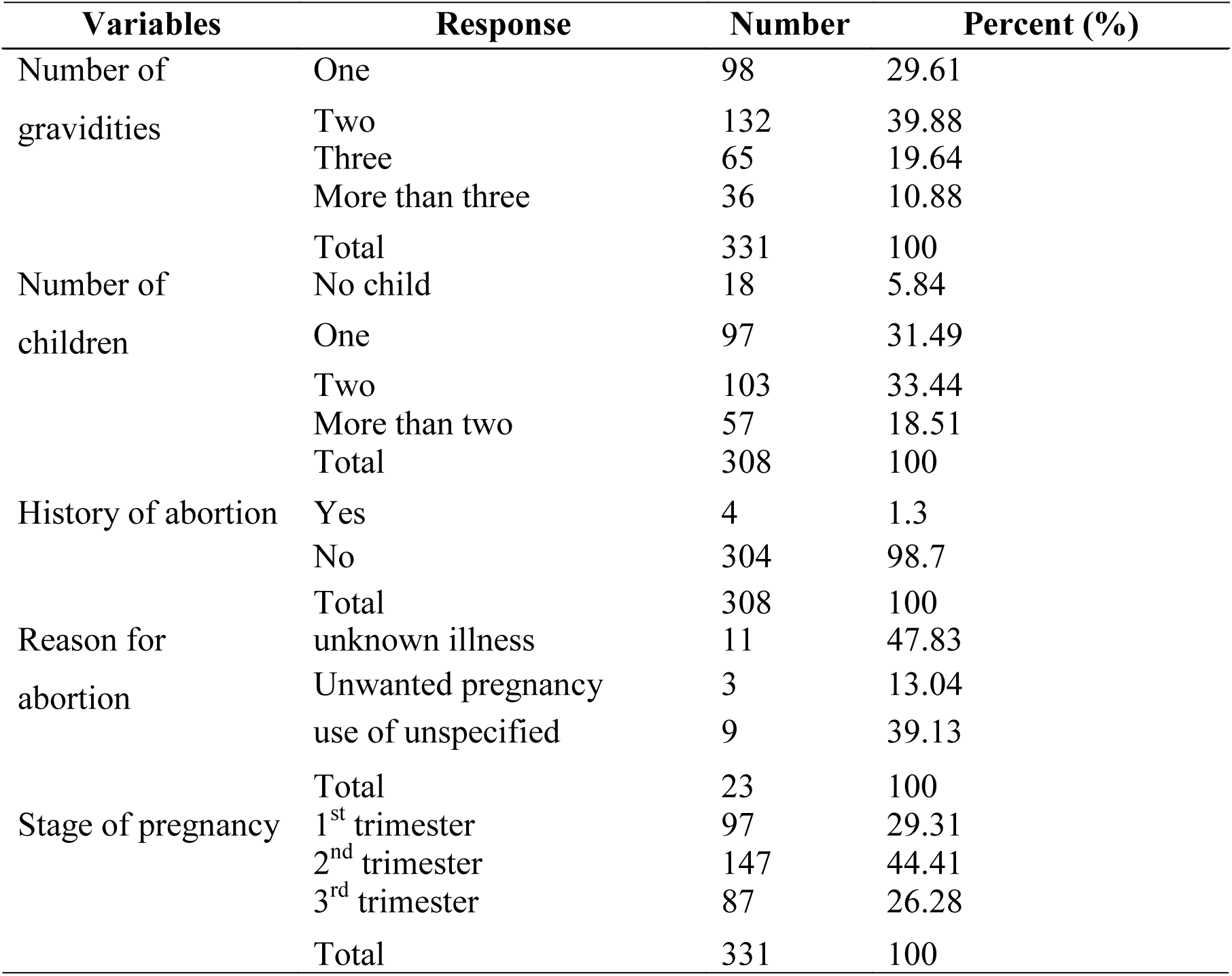
Obstetric of pregnant women attending ANC at JUMC

### Self-medication use of herbal medicine

Thirteen (3.93%) of the 331 pregnant women who were questioned for this study currently self-medicate, while none (0.00%) of the 331 pregnant women overall have ever self-medicated in the past (Table 3). Ginger (1.5%) and “damakasse” (1.21%) were the two herbs that the respondent used most frequently. According to the study’s findings, herbal medication was frequently used to treat headaches (1.21%) and vomiting (1.81%). Family and friends (2.11%) and traditional healers (1.21%) were the most popular sources of knowledge for self-medication among pregnant women who used it during their most recent pregnancy (1.21) (Table 3).

**Table 3.** Self-medication of herbal medicine among pregnant women attending ANC at JUMC

| Variables | Response | Number | Percent (%) |
| --- | --- | --- | --- |
| Herbal medicine used during a prior pregnancy | Yes | 0 | 0.00 |
|  | No | 331 | 100.00 |
| Herbal medicine used during the current pregnancy | Yes | 13 | 3.93 |
|  | No | 318 | 96.07 |
| Reason for practicing herbal medicine | For its effectiveness | 0 | 0.00 |
|  | For its fewer side effects | 0 | 0.00 |
|  | For its cheapest | 0 | 0.00 |
|  | For its accessibility | 13 | 100.00 |
| The purpose of the use of herbal medicine | During headache | 4 | 1.21 |
|  | During nausea/vomiting | 6 | 1.81 |
|  | During typhoid | 3 | 0.91 |
|  | During UTI | 0 | 0.00 |
|  | During Common cold | 0 | 0.00 |
|  | During diarrhea | 0 | 0.00 |
|  | To facilitate labor | 0 | 0.00 |
|  | To prevent abortion | 0 | 0.00 |
| Type of herbs used | Ginger | 5 | 1.51 |
|  | Garlic | 3 | 0.91 |
|  | Tenaadam | 1 | 0.30 |
|  | Damakasse | 4 | 1.21 |
|  | Tosign | 0 | 0.00 |
|  | Other/specify | 0 | 0.00 |
| The information source of herbal medicine used | Traditional healers | 4 | 1.21 |
|  | Health professionals | 0 | 0.00 |
|  | Religious leaders | 0 | 0.00 |
|  | Family and friends | 7 | 2.11 |
|  | Neighbors | 2 | 0.60 |
|  | Other/specify | 0 | 0.00 |

## Discussions

Regardless of self-medications, only 3.93% of 331 pregnant women who were questioned for this study currently self-medicate. This was significantly lower than that of the study conducted in Ghana in which the prevalence of self-medication practice was 74.1% [11]. Another study done in Tanzania revealed that the prevalence of self-medication among pregnant women was 172 (46.24%). This might be due to a lack of awareness of the self-medication practices in our recent study. Pregnant women with no history of self-medication become self-medicated currently. This finding was also different from the study done in Mizan Tepi University Teaching Hospitals in which women with no history of self-medication were 6.69 folds less likely to practice self-medication than those with prior experience [12]. The reason for this might be because of a lack of exposure to self-medication to understand self-medication impact.

The most frequently used herbal medicine by pregnant women in our study were ginger (1.5%) and damakasse (1.21%). This finding was almost different from the study done in tertiary hospitals of Jimma in which the most commonly used medicinal plants were Linum usitatissimum L. (flaxseed—use with caution) 22.0%, Ocimum lamiifolium L. (damakesse— safety unknown) 3.6% and Carica papaya L. (papaya—use with caution) 3.1% [13]. This may result from the easy availability of those herbal medicine used in the environment in our cases.

Almost all of the users of herbal medicines were due it its easy accessibility which was similar to the study conducted in Kemise General Hospital in which the majority of participants practiced herbal medicine for the reason of being easily accessible [14]. The most common purpose for the use of herbal medicine by pregnant women attending antenatal care in our case was vomiting and headache respectively. But the study conducted in Shashemene town revealed that constipation and headaches were the most common indication of herbal medicine respectively [15]. The possible reason for this might be the prevalence of indications were different from geographic location to geographic location as well as the presence of different attitude among the community. A systematic review conducted in Ethiopia shows the herbal medicines ginger and damakasse are commonly consumed by women during pregnancy which was similar to our study [6].

The most common source of information for herbal medicines are family and friends which was different from the study done in the Gedeo zone in which the source of information was a marketplace [16]. The rationale for our case might be living together most of the time.

### Limitations of study

The current study did not evaluate the impact of herbal medicine on pregnant women and infants during birth among those who practiced herbal medicine use. Another limitation of this study was its cross-sectional nature to determine causal relationship.

## Conclusions

Self-medication practice of herbal medicine prevalence was 3.93% during their current pregnancy. The most frequently practiced herbal medicine among pregnant women attending antenatal care at JUMC were Ginger and Damakese with the most common clinical indication of vomiting and headache respectively. The Source of information for the participant’s herbal medicine use was commonly family and friends.

## Data Availability

Data sets are available

## List of acronyms

ANC: Antenatal care
ETB: Ethiopian Birr
IBR: Institutional Review Board
JUMC: Jimma University Medical Center
POMs: Prescription only Medications
SSA: Sub-Saharan Africa
TM: Traditional Medicine
WHO: World Health Organizations.

## Acknowledgments

Not applicable

## Authors contribution

Firafan Shuma: Conceptualization; Data Curation; Investigation; Methodology; Validation; Writing – Original Draft Preparation; Writing – Review & Editing. Damisie Beharu: Conceptualization; Data Curation; Investigation; Methodology; Validation; Writing – Original Draft Preparation; Writing – Review & Editing. Dugasa Erko: Data Curation; Investigation; Methodology; Validation; Writing – Original Draft Preparation. Tamirat Tekassa: Data curation; Validation; Writing-Original Preparations. All authors read and approved the final manuscript.

## Patient consent for publication

Not applicable

## Ethical approval

This study was approved by the Jimma University Institute of Health Institutional Review Board (IRB) (ref. no. JUIH 143422/22) in Ethiopia

## Data availability statement

The data used for analysis can be provided upon a request to the corresponding author

